# Neutralization profile of Omicron variant convalescent individuals

**DOI:** 10.1101/2022.02.01.22270263

**Authors:** Annika Rössler, Ludwig Knabl, Dorothee von Laer, Janine Kimpel

## Abstract

Recently, the Omicron variant of the severe acute respiratory syndrome coronavirus 2 (SARS-CoV-2) has been described as immune escape variant.

Here, we analyzed samples from BA.1 (Omicron) convalescent patients with different constellations of prior SARS-CoV-2 immunity regarding vaccination and previous infection with a non-Omicron variant and determined titers of neutralizing antibodies against different SARS-CoV-2 variants (D614G, Alpha, Beta, Delta, Gamma, Omicron).

We found high neutralizing antibody titers against all variants for vaccinated individuals after BA.1 breakthrough infection or for individuals after infection with a pre-omicron variant followed by BA.1 infection. In contrast, samples from naive unvaccinated individuals after BA.1 infection mainly contained neutralizing antibodies against BA.1 but only occasionally against the other variants.

---

We and others have previously shown that sera from unvaccinated individuals after Alpha (B.1.1.7), Beta (B.1.351) or Delta (B.1.617.2) variant infection only occasionally neutralize the Omicron variant.(1) Similarly, Omicron neutralizing antibodies are low and only short lived after one or two doses of CODID-19 vaccination, but enhanced in hybrid immune individuals (combination of vaccination and infection) or after a third booster dose of vaccination.(2, 3)

Only little is known about neutralization profiles of Omicron variant convalescent individuals.(4, 5) So far, either vaccinated individuals after Omicron breakthrough infections or unvaccinated individuals, where the history of previous infection is unknown, were analyzed. Therefore, we analyzed here neutralization profiles against six different SARS-CoV-2 variants for plasma samples from Omicron (BA.1) convalescent individuals with four different combinations of prior SARS-CoV-2 immunity regarding vaccination and previous infection with a non-Omicron variant.

We collected plasma samples from individuals 5-35 days after the first positive PCR during BA.1 infection. We included individuals with four different constellations of SARS-CoV-2 immunity prior to BA.1 infection; (i) vaccinated without prior history of infection (n=15), (ii) unvaccinated without prior history of infection (n=13), (iii) vaccinated with prior infection (n=10), and (iv) unvaccinated with prior infection (n=13), for details regarding patient characteristics and vaccination regiments see Supplementary Appendix Tables S1-S4. We analyzed neutralizing antibodies using a panel of replication competent SARS-CoV-2 variants including parental D614G, B.1.1.7, B.1.351, P1.1 (Gamma), B.1.617.2, and BA.1 variants.

We found high neutralizing antibody titers against all variants for vaccinated individuals after BA.1 breakthrough infection or for individuals after infection with a pre-omicron variant followed by BA.1 infection (Figure 1). Interestingly, BA.1 neutralizing antibody titers were reduced compared to titers to other variants in vaccinated individuals but comparable in unvaccinated individuals with D614G or B.1.617.2 infection followed by BA.1 infection. In contrast, samples from naive unvaccinated individuals after BA.1 infection mainly contained neutralizing antibodies against BA.1 but only occasionally against the other variants. Surprisingly, two unvaccinated individual had only neutralizing antibodies against the BA.1 and P.1.1 variant despite a single previously positive PCR not accompanied by any symptoms for B.1.617.2 variant 2 months prior to re-infection (Figure 1D). As these individuals did not neutralized B.1.617.2 nor most other variants apart from BA.1, the previous B.1.617.2 PCR might have been false-positive.

**Figure 1.**
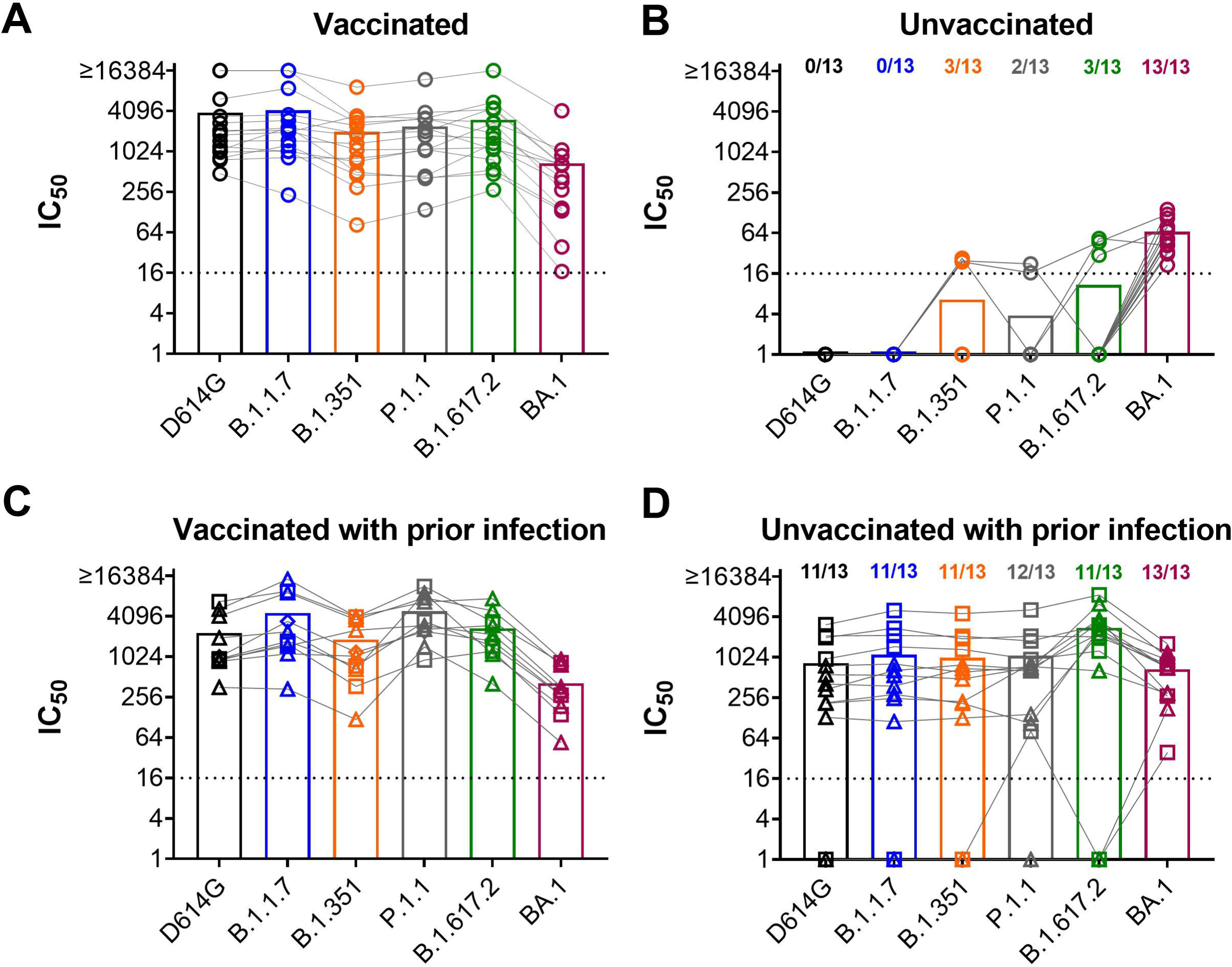
Neutralization capacity of plasma samples from BA.1 (Omicron) convalescent patients. Plasma samples were collected from BA.1 convalescent patients; Panel A vaccinated individuals with no prior infection (n=15), Panel B unvaccinated individuals with no prior infection (n=13), Panel C vaccinated individuals with a prior history of D614G (squares), B.1.1.7 (diamonds), or B.1.617.2 (Delta variant, triangles) SARS-CoV-2 infection (n=10), Panel D unvaccinated individuals with a prior history of either D614G (squares) or B.1.617.2 (triangles) SARS-CoV-2 infection (n=13). Plasma was collected 5-35 days after first positive PCR (BA.1 infection). Samples were analyzed for 50 % neutralization titers (IC50) using life D614G, B.1.1.7 (Alpha), B.1.351 (Beta), P.1.1 (Gamma), B.1.617.2 (Delta), or BA.1 SARS-CoV-2 isolates. Individual values and mean titers (bars) are shown. Samples for each individual patient analyzed with the different virus variants are connected by lines. Titers below 1:16 are regarded as negative (dotted line).

Our data support the hypothesis that the Omicron variant represents a distinct new serotype. Therefore, unvaccinated individuals, infected with the Omicron variant only, might not be protected well against infection with a non-Omicron SARS-CoV-2 and should consequently be vaccinated in addition for full protection.

## Data Availability

All data produced in the present study are available upon reasonable request to the authors.

