## Supplementary Appendix for "Neutralization profile of Omicron variant convalescent individuals"

### List of Investigators

Annika Rössler[1]

Ludwig Knabl[2]

Dorothee von Laer[1]

Janine Kimpel[1]

[1]Institute of Virology, Department of Hygiene, Microbiology and Public Health, Medical University of Innsbruck, Peter-Mayr-Str. 4b, 6020 Innsbruck, Austria

[2]Tyrolpath Obrist Brunhuber GmbH, Hauptplatz 4, 6511, Zams, Austria

#### Methods

*Ethics statement*

The ethics committee (EC) of the Medical University of Innsbruck has approved the study with EC numbers 1064/2021 and 1191/2021.

*Plasma samples*

Plasma samples were collected 5 to 35 days after confirmed diagnosis of BA.1 infection by qPCR. Of the unvaccinated cohort, 4 individuals had a previous infection with D614G and 9 individuals had been tested positive for B.1.617.2 one to 24 months before their reinfection with BA.1. In addition, 3 and 6 individuals in the vaccinated group had a confirmed prior infection with D614G or B.1.617.2 respectively one to 22 months before reinfection with BA.1. Moreover, one participant had tested positive for B.1.1.7 ten months before reinfection with BA.1.

*Focus forming neutralization assay*

An adapted protocol of a previously described [1] focus forming neutralization assays was performed using replication competent SARS-CoV-2 isolates (D614G: Isolate B86.2, GISAID ID EPI_ISL_3305837; B.1.1.7: isolate C69.1, GISAID ID EPI_ISL_3277382; B.1.351: isolate C24.1, GISAID ID EPI_ISL_1123262; P1.1: isolate hCoV-19/Germany/BY-MVP-000005870/2021, GISAID ID EPI_ISL_2095177; B.1.617.2: isolate SARS-CoV-2-hCoV-19/USA/NY-MSHSPSP-PV29995/2021, GISAID ID EPI_ISL_2290769; BA.1: isolate E16.1, GISAID ID EPI_ISL_6902053;). Briefly, plasma samples were heat inactivated for 30 minutes at 56 °C and clarified by centrifugation at 8,000 rpm for 5 minutes. Viruses were incubated in duplicates with serially four-fold diluted samples (1:16 to 1:16,384 dilutions in complete medium with 2% fetal calf serum) for 1 hour at 37 °C. Subsequently, confluent Vero cells stably expressing TMPRSS2 and ACE2 receptor were inoculated with serum/virus mixtures for 2h, resulting in approximately 100-200 infected cells in control wells without serum. Thereafter, serum/virus mixes were replaced by fresh medium and 8 hours later cells were fixed for 5 minutes using 96% ethanol. Infected cells were stained using serum of a convalescent patient and an Alexa Fluor Plus 488-conjugated goat anti-human IgG secondary antibody (Invitrogen, Thermo Fisher Scientific, Vienna, Austria). Infected cells were counted using an ImmunoSpot S6 Ultra-V reader and FluoroSpot software (CTL Europe GmbH, Bonn, Germany). Continuous 50% neutralization titers were calculated in GraphPad Prism 9.0.1 (GraphPad Software, Inc., La Jolla, CA, USA) using a non-linear regression. Titers <1:16 were considered negative. Titers >1:16,384 were set to 1:16,384.

*Statistics*

One-Way paired ANOVA (Friedman's test) with Dunn's multiple comparisons was used to determine statistical differences using GraphPad Prism 9.0.1 (GraphPad Software, Inc., La Jolla, CA, USA).

*Acknowledgments*

We thank Albert Falch, Eva Hochmuth, Evelyn Peer, Lisa-Maria Raschbichler, Bianca Neurauter, Lydia Riepler, David Bante, Lukas Perro, Stephan Amstler, Andreas Aufschnaiter, Luiza Hoch, Helena Schäfer for excellent technical and organizational support. We thank Prof. Florian Krammer and Prof. Viviana Simon for sharing their Delta isolate and Prof. Oliver T. Keppler and Dr. Marcel Stern for sharing their Gamma isolate with us.

#### Table S1. Patient characteristics of vaccinated individuals without prior history of pre-Omicron infection

| AGE (YEARS) | SEX[#] | DAYS SINCE BA.1 INFECTION* | VACCINATION |
|---|---|---|---|
| **<20** | f | 10 | BNT162b/BNT162b |
| **50-59** | m | 18 | BNT162b/BNT162b |
| **50-9** | f | 18 | BNT162b/BNT162b |
| **30-39** | m | 20 | BNT162b/BNT162b |
| **30-39** | m | 8 | BNT162b/BNT162b/BNT162b |
| **20-29** | m | 14 | BNT162b/BNT162b/BNT162b[%] |
| **20-29** | m | 33 | ChAdOx1 |
| **20-29** | f | 35 | BNT162b |
| **30-39** | m | 23 | mRNA-1273/mRNA-1273/mRNA-1273[%] |
| **20-29** | f | 10 | BNT162b/BNT162b |
| **30-39** | f | 6 | mRNA-1273/mRNA-1273/BNT162b[%] |
| **20-29** | f | 10 | BNT162b/BNT162b |
| **20-29** | f | 5 | Ad26.COV2.S/BNT162b |
| **80-89** | m | 9 | BNT162b/BNT162b/BNT162b |
| **40-49** | f | 12 | ChAdOx1/ChAdOx1 |

### f = female; m = male; * Days since first positive PCR for BA.1 infection; [%]Interval between last dose of vaccination and first positive PCR for BA.1 infection less than 14 days

#### Table S2. Patient characteristics of unvaccinated individuals without prior history of pre-Omicron infection

| AGE (YEARS) | SEX[#] | DAYS SINCE BA.1 INFECTION* |
|---|---|---|
| **<20** | m | 13 |
| **40-49** | f | 17 |
| **40-49** | m | 14 |
| **60-69** | m | 14 |
| **30-39** | m | 14 |
| **30-39** | f | 14 |
| **50-59** | f | 14 |
| **50-59** | m | 14 |
| **20-29** | f | 11 |
| **20-29** | m | 11 |
| **60-69** | f | 12 |
| **80-89** | f | 12 |
| **40-49** | f | 16 |

### f = female; m = male; * Days since first positive PCR for BA.1 infection

#### Table S3. Patient characteristics of vaccinated individuals with prior history of pre-Omicron infection

| AGE (YEARS) | SEX# | DAYS SINCE BA.1 INFECTION* | VACCINATION | PRIOR INFECTION | |
|---|---|---|---|---|---|
| | | | | MONTHS§ | VARIANT |
| **50-59** | m | 12 | BNT162b/BNT162b | 7 | B.1.617.2 |
| **20-29** | f | 13 | ChAdOx1/ChAdOx1 | 2 | B.1.617.2 |
| **40-49** | m | 12 | BNT162b | 13 | D614G |
| **20-29** | m | 13 | BNT162b | 2 | B.1.617.2 |
| **40-49** | m | 13 | ChAdOx1/ChAdOx1/BNT162b | 2 | B.1.617.2 |
| **50-59** | f | 16 | ChAdOx1/ChAdOx1/BNT162b | 2 | B.1.617.2 |
| **30-39** | f | 13 | BNT162b/BNT162b | 1 | B.1.617.2 |
| **20-29** | f | 10 | BNT162b/BNT162b | 14 | D614G |
| **20-29** | m | 19 | BNT162b | 10 | B.1.1.7 |
| **50-59** | f | 27 | BNT162b/ChAdOx1 | 22 | D614G |

### f = female; m = male; * Days since first positive PCR for BA.1 infection; § Months since confirmed diagnosis of prior D614G or B.1.617.2 infection by qPCR

#### Table S4. Patient characteristics of unvaccinated individuals with prior history of pre-Omicron infection

| AGE (YEARS) | SEX# | DAYS SINCE BA.1 INFECTION* | PRIOR INFECTION | |
|---|---|---|---|---|
| | | | MONTHS§ | VARIANT |
| **<20** | f | 13 | 21 | D614G |
| **50-59** | m | 17 | 21 | D614G |
| **50-59** | f | 14 | 14 | D614G |
| **20-29** | f | 7 | 5 | B.1.617.2 |
| **30-39** | m | 10 | 1 | B.1.617.2 |
| **<20** | f | 8 | 2 | B.1.617.2 |
| **<20** | f | 14 | 2 | B.1.617.2 |
| **<20** | m | 14 | 2 | B.1.617.2 |
| **20-29** | m | 15 | 1 | B.1.617.2 |
| **60-69** | m | 13 | 2 | B.1.617.2 |
| **<20** | f | 15 | 2 | B.1.617.2 |
| **40-49** | m | 18 | 14 | D614G |
| **50-59** | f | 11 | 2 | B.1.617.2 |

### f = female; m = male; * Days since first positive PCR for BA.1 infection; § Months since confirmed diagnosis of prior D614G or B.1.617.2 infection by qPCR

#### Table S5. Statistics

| | D614G# | B.1.1.7# | B.1.351# | P.1.1# | B.1.617.2# |
|---|---|---|---|---|---|
| **VACCINATED** | **** | **** | ns | ** | **** |
| **UNVACCINATED** | *** | *** | ** | *** | ** |
| **VACCINATED WITH PRIOR INFECTION** | ns | **** | * | **** | ** |
| **UNVACCINATED WITH PRIOR INFECTION** | ns | ns | ns | ns | ns |

#Statistical difference to BA.1 using ANOVA, followed by Friendman`s test with Dunn's multiple comparisons; * $p<0.05$; ** $p<0.01$; *** $p<0.001$; **** $p<0.0001$; ns = non-significant;
